# Biological Aging for Risk Prediction of First-Ever Intracerebral Hemorrhage and Cerebral Infarction in Advanced Age

**DOI:** 10.1101/2021.10.28.21265620

**Authors:** Reem Waziry, Albert Hofman, Mohsen Ghanbari, Henning Tiemeier, M. Arfan Ikram, Anand Viswanathan, Jaco Klap, M. Kamran Ikram, Jaap Goudsmit

## Abstract

Successful interventions to prevent cerebrovascular disease and stroke require early identification of persons at risk before clinical manifestation of disease. We assessed the predictive value of biological age (BA) as an early indicator for cerebrovascular disease and risk of first-ever intracerebral hemorrhage (ICH) and cerebral infarction (CI) in advanced age and compared these relationships with commonly used biomarkers including tau and Aβ40 and Aβ42. The study included Individuals who consented for blood draw and follow-up. We computed biological age using structural equation modelling. The algorithm integrates biomarkers that represent six body systems involved in overall cerebrovascular health including metabolic function, cardiac function, lung function, kidney function, liver function, immunity and inflammation. Time to event analysis was conducted using Cox-regression models. Prediction analysis was conducted using Harrel’s C and Area under the receiver operating characteristic curve. The sample included a total of 1699 individuals at baseline followed up over a median of 11 years. During a period of 15, 780 and 16, 172 person-years a total of 17 first-ever intracerebral hemorrhage and 83 cerebral infarction cases occurred. In time-to-event analysis, BA showed higher magnitude of associations with ICH compared to CA (HR_BA-ICH:_ 2.30, 95% CI: 1.20, 4.30; HR_CA-ICH:_ 1.40, 95% CI: 0.76, 2.53) and higher precision with CI (HR_BA-CI:_ 1.30, 95% CI: 1.01,1.75; HR_CA-CI:_1.90, 95% CI: 1.48, 2.66). BA outperformed CA for prediction of ICH (AUC: 0.68 vs 0.53; Harrel’s C: 0.72 vs 0.53) and for CI (AUC:0.63 vs 0.62; Harrel’s C: 0.68 vs 0.67). Biological aging based on integrated physiology biomarkers provides a novel tool for monitoring and identification of persons at highest risk of cerebrovascular disease in advanced age. Future studies should confirm these observations in bigger samples and further characterize aging mechanisms that contribute towards brain reserve and resilience among individuals with similar risk profiles.

## INTRODUCTION

The risk of intracerebral hemorrhage (ICH) and cerebral infarction (CI) double with every decade of life and continue to escalate with the aging of populations ^1, 2^. Chronological age is not a specific indicator for risk prediction of cerebrovascular disease. Data from autopsy studies have shown wide variability between individuals in their ability to tolerate clinically occult pathological changes and physiological degradation before manifestation of disease ^3, 4^. Traditional approaches that account for aging through chronological age overlook within and between individual variations in aging, that are hypothesized to be root causes of these devastating disorders ^5^. These observations collectively suggest that variations between individuals might be linked to individual specific factors such as life-time exposures and resiliency ^6, 7^.

The relationship between biological aging using integrated physiology approaches and risk of incident (new-onset) age-related cerebrovascular disorders has not been assessed before. Previous studies that assessed relationship between aging biomarkers and health outcomes were limited in design and suffered temporality in assessment of disease endpoints. The especial emphasis on physiology-based biomarker panels is informed by previous studies that observed weak to moderate associations of other biological aging metrics with health outcomes beyond what is explained by chronological age ^8-10^.

In the present study, we assessed the relationship between biological age and risk of first occurrence of intracerebral hemorrhage and cerebral infarction in advanced age and compared these relationships with commonly used biomarkers including tau and Aβ40 and Aβ 42.

## METHODS

### The Rotterdam Study

The present report included participants from the Rotterdam Study, a prospective population-based cohort ^11, 12^. In 1990, residents aged 55 years and older residing in Ommoord, a district of Rotterdam, the Netherlands, were invited to participate. Of 10,215 invited inhabitants, 7,983 agreed to participate in the baseline examinations. In 2000, 3,011 participants (of 4,472 invitees) who had reached 55 years of age or moved into the study district since the start of the study were added to the cohort. Follow-up examinations take place every 3–4 years. For the purpose of this study, 2000 individuals were randomly selected from the fourth visit of Rotterdam Study-I (RS-I-4) and the second visit of Rotterdam Study-II (RS-II-2). The inclusion criteria included the availability of informed consent and valid serum samples collected between 2002 and 2005 in these visits of Rotterdam study I and II (Figure 1, Supplementary Figure 1).

**Figure 1.**
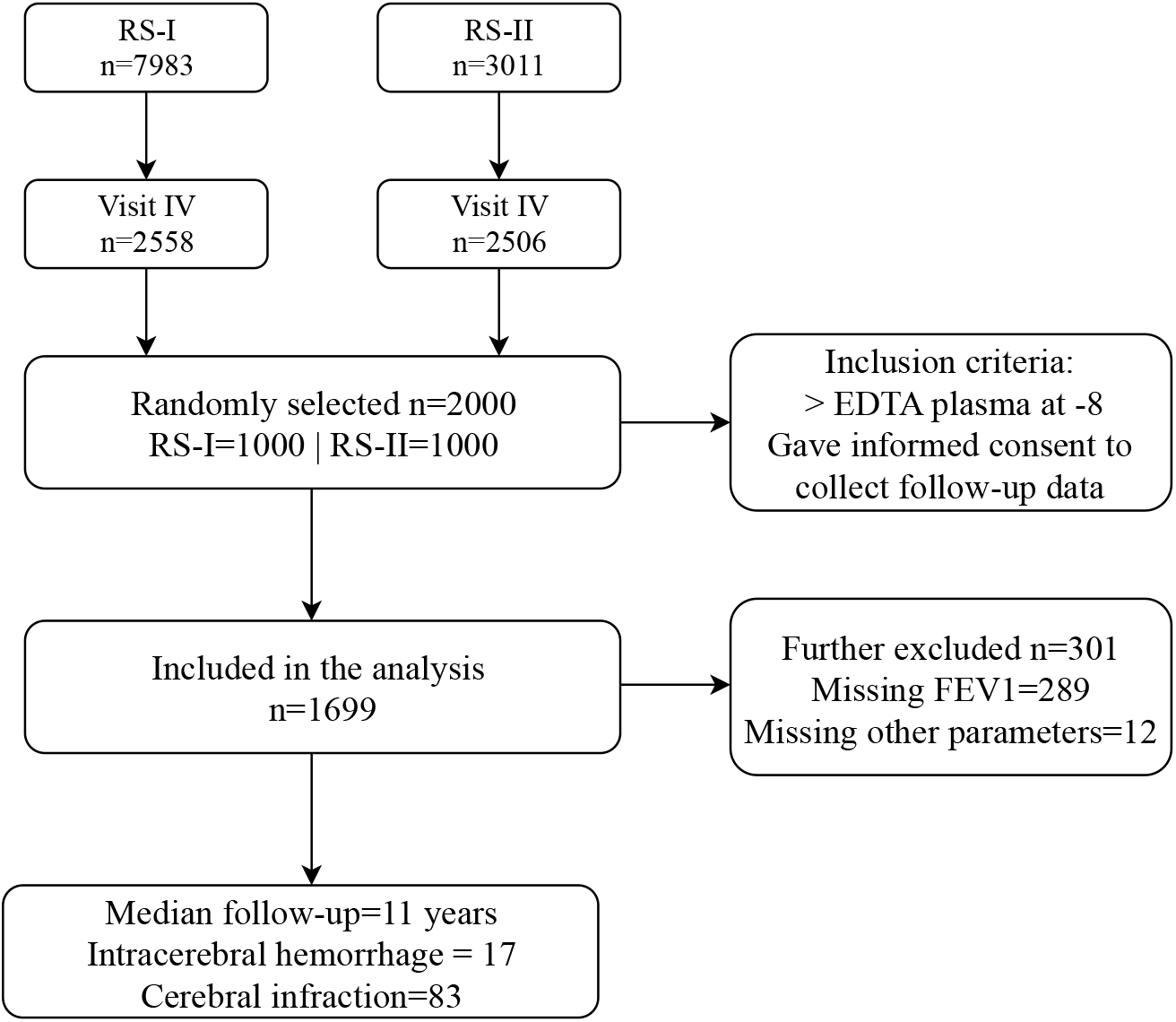
Study flowchart.

**Figure 2.**
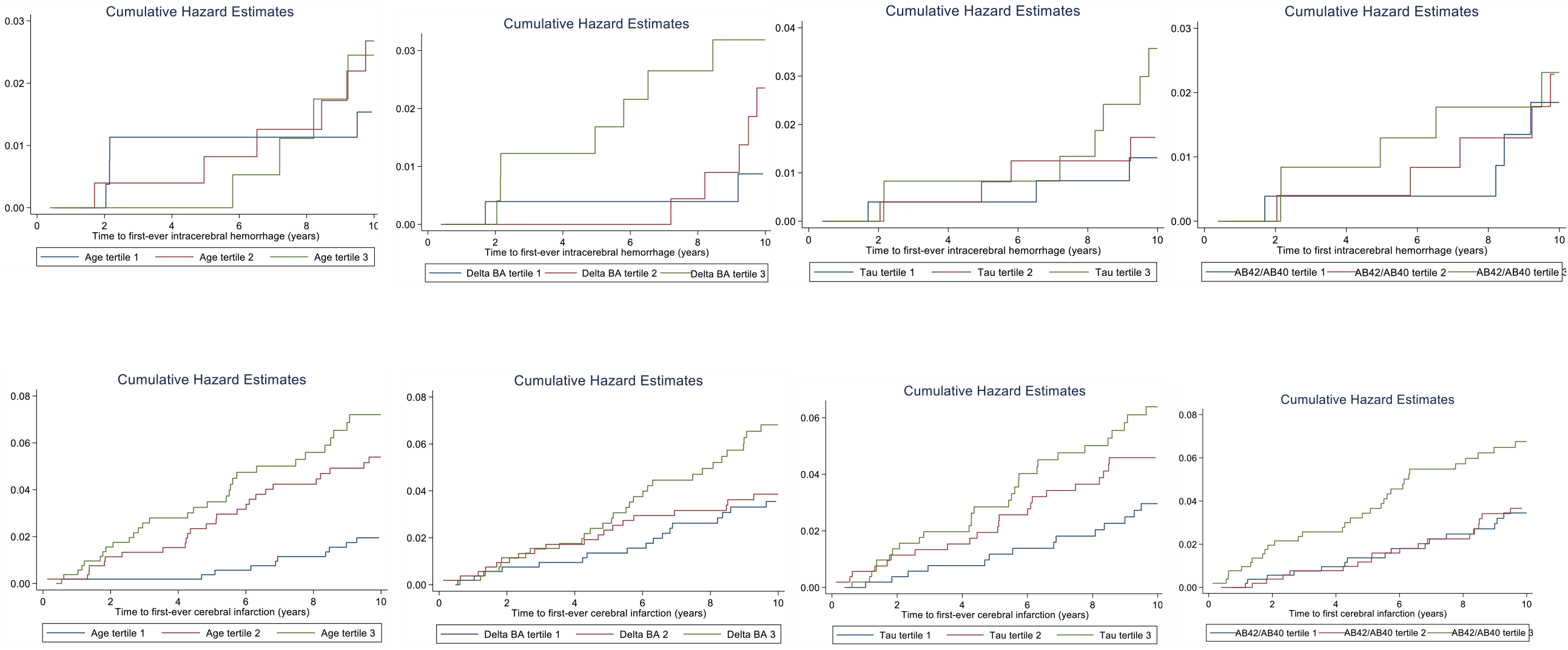
Cumulative hazards of time to intracerebral hemorrhage (upper row) or cerebral infarction (lower row) by tertiles of biomarkers.

### Informed Consent and Ethics Approval

The Rotterdam Study has been approved by the medical ethics committee of the Erasmus MC (registration number MEC 02.1015) and by the Dutch Ministry of Health, Welfare and Sport (Population Screening Act WBO, license number 1071272-159521-PG). When visiting the study center, participants provided written informed consent to participate in the study and to have their information obtained from treating physicians.

### NHANES

The reference population included participants from the Third National Health and Nutrition Examination Survey (NHANES III), a nationally representative, cross-sectional study conducted by the National Center for Health Statistics conducted between 1988 and 1994. Data collection was done through at-home interviews and examinations. Further details on study population and design are available through the Centers for Disease Control and Prevention ^13^. This external independent population was used to train and calibrate the biological aging algorithm to be assessed for risk prediction of our outcomes of interest in the Rotterdam Study population.

### Stroke ascertainment in the Rotterdam study population

Stroke was defined according to the World Health Organization criteria as a syndrome of rapidly developing clinical signs of focal (or global) disturbance of cerebral function, with symptoms lasting 24 hours or longer or leading to death, with no apparent cause other than of vascular origin ^12, 14, 15^. History of stroke at baseline was assessed during baseline interview and verified by reviewing medical records. After enrollment, participants were continuously monitored for incident stroke through automated linkage of the study database with files from general practitioners. Nursing home physicians’ and general practitioner files of participants who moved out of the district were checked on a regular basis as well. Additional information was obtained from hospital records. The information collected on potential strokes was reviewed and classified by research physicians and verified in a consensus panel led by two experienced stroke neurologists. Final stroke diagnosis was adjudicated in accordance with the above mentioned standardized diagnostic WHO criteria, which were held constant over the entire follow-time. Strokes were further classified as ischemic or primary intracerebral haemorrhage based on neuroimaging reports or hospital discharge letters. If neuroimaging was not conducted or reported, a stroke was classified as unspecified. This classification corresponds with ICD-10 codes I61, I63 and I64. In the stroke analysis, participants could contribute to follow-up until first-ever stroke, death, loss to follow-up or last health status update when they were known to be free of stroke, whichever came first, until January 2016. Follow-up was virtually complete (97.8%).

### Quantification of biological age

A detailed description of the biological aging calculation is provided in a previous report ^16^. Briefly, nine biomarkers were selected based on: 1) their independent relationship with age; 2) their availability in the Rotterdam Study, and; 3) validation of the algorithms in previous studies ^17, 18^. The biomarkers represent six systems as follows: total cholesterol (metabolic function); systolic blood pressure (cardiac function); forced expiratory volume (lung function); serum creatinine and serum urea nitrogen (kidney function); serum alkaline phosphatase and serum albumin (liver function) and C-reactive protein, cytomegalovirus optical density (immune function and inflammation) ^16^. These biomarkers were measured using serum. (Table 1, Figure 1, Supplementary Figures 1, Supplementary Tables 1-3).

**Table 1.** Population characteristics.

|  | **All** | **Intracerebral**<br>**hemorrhage** | **Cerebral infarction** |
| --- | --- | --- | --- |
| N | 1699 | 17 | 83 |
| Age, baseline | 70 (65, 76) | 71 (66, 76) | 74 (70, 79) |
| Female | 967 (57) | 8 (47) | 61 (73) |
| BMI, mean (SD) | 27 (25, 30) | 26 (24, 27) | 27 (25, 29) |
| BMI categories<sup>a</sup> |  |  |  |
| ≤25 | 464 (27) | 8 (47) | 29 (35) |
| 25-30 | 806 (48) | 9 (52) | 66 (79) |
| >30 | 421 (25) | 3 (17) | 24 (29) |
| APOE4% | 430 (25) | 7 (41) | 31 (37) |
| **Individual biomarkers<sup>b</sup>** |  |  |  |
| *C-reactive protein (mg/dL)* | 0.30 (0.10–15) | 0.21 (0.10–0.40) | 0.21 (0.10–0.50) |
| *Creatinine (mg/dL)* | 0.90 (0.50–40) | 0.96 (0.91–1.04) | 0.98 (0.84–1.14) |
| *Albumin (g/dL)* | 4.00 (3.00–5.00) | 4.60 (4.40–4.70) | 4.50 (4.30–4.60) |
| *Total cholesterol (mg/dL)* | 217 (86–360) | 207 (177–241) | 209 (178–235) |
| *Cytomegalovirus optical density* | 87 (5–180) | 1.80 (1.20–2.10) | 1.90 (0.70–2.10) |
| *Urea nitrogen (mg/dL)* | 15 (4–66) | 15.90 (12.70–19.60) | 15.90 (13.40–19.80) |
| *Alkaline phosphatase SI (U/L)* | 78 (22–437) | 77.5 (72.50–95.50) | 79 (65–95) |
| *Forced expiratory volume (mL)* | 2 (0.20–7.20) | 2.30 (1.80–3.10) | 2.20 (1.80–2.90) |
| *Systolic blood pressure* | 146 (92–225) | 161.5 (139.5–172.7) | 150.5 (136.5–164.5) |

BMI was categorized according to the CDC definition of normal (18.5-25), overweight; 25-30; obese 30 or higher. b Individual biomarkers values rounded to first decimal place

Biological age was calculated as the weighted mean of these 9 parameters using structural equation modelling (Supplementary Figure 5). Two separate models were used for men and women. The calculations included three main steps in SAS software. First, we used the function PROC CALIS to generate a set of weights (i.e., coefficients) using biomarker data from NHANES for multiplication with chronological age and the biomarkers in the structural equation models ^19^. These weights are the product of the variance covariance matrix ^20^. Second, we multiplied the biomarker weights by chronological age and the biomarker values through PROC SCORE. Finally, we scaled the calculated biological age to the chronological age scale. This is an alternative to normalizing chronological age and biomarkers using the mean and standard deviation. Given biomarkers of whole blood samples (as HBA1c) were not available in our data, we compared nine versus ten biomarkers algorithm in NHANES and the results showed similar results of hazard ratios and model performance based on Akaike Information Criterion (AIC) and Bayesian Information Criterion (BIC) (Supplementary Table 3).

### Measurement of biomarkers of total-tau, Aβ 40 and Aβ 42

EDTA plasma was sampled, aliquoted and frozen at -80°C according to standard procedures. Measurements were obtained from a random selection of 1000 participants from sub-cohort RS-I-4 and 1000 from RS-II-2. All measurements were performed at Quanterix (Lexington, MA, USA) on a Single Molecule Array (Simoa) HD-1 analyzer platform. Samples were tested in duplicate. Two quality control (QC) samples were run on each plate for each analyte. The Simoa Human Neurology 3-Plex A assay (N3PA) was used for measuring the concentration of total-tau, Aβ 40 and Aβ 42. When duplicates or single measurements were missing or in the case the concentration coefficient of variation (CV) exceeded 20% or control samples were out of range, participant’s data were excluded from the analyses. A dedicated investigation of biomarkers of neurodegeneration in relation to stroke has been conducted in a larger sample of the Rotterdam Study^27^.

### Statistical analysis

Delta biological age was calculated as the difference between biological age and chronological age for each individual in the Rotterdam Study population. Delta biological age in this case represents the absolute difference between predicted biological age and observed chronological age. We assessed onset of two clinical endpoints: 1) Intracerebral hemorrhage; and 2) Cerebral infarction. Follow-up was defined as time from visit date (date of biological age measurement) at which biological age was calculated to stroke date, death or end of follow-up on January 1^st^, 2016, whichever occurred first. In the time to event analysis using Cox model, individuals who had stroke date equals death date or end of study follow-up, contributed one day of follow-up in the analysis ^21^. Those with prevalent stroke or unspecified stroke diagnosis were excluded. Models were adjusted for age, sex and BMI. Biomarkers were categorized in tertiles and assessed for risk prediction of intracerebral hemorrhage and cerebral infarction in separate models. Area under the receiver operating characteristic curve (AUC) analysis was conducted to assess the sensitivity (true positive rate) and specificity (true false rate) of delta biological age for prediction of intracerebral hemorrhage and cerebral infarction.

## RESULTS

### Population characteristics

The sample included a total of 1699 individuals at baseline followed up over a median of 11 years. During a total of 15, 780 person-years a total of 17 intracerebral hemorrhage cases occurred, among which 47% were females, 52% were overweight and 41% were APOE4 carriers. During 16, 172 person-years a total of 83 cerebral infarction cases occurred, among which 73% were females, 79% were overweight and 37% were APOE4 carriers (Table 1).

### Biological age as a predictor of risk of ICH and IC

In time-to-event analysis risk of intracerebral hemorrhage (compared to the lowest tertile) was 2.30 (95% CI 1.20, 4.30) per tertile increase of delta biological age. We compared this in the same sample for which our biological aging biomarkers are computed to per tertile change in chronological age 1.40 (95% 0.76, 2.53). BA exceeded CA for prediction of ICH (AUC: 0.68 vs 0.53; Harrel’s C: 0.72 vs 0.53) (Tables 1-2, Figure 1, Supplementary figures 2-4, Supplementary table 1).

In time-to-event analysis, risk of cerebral infarction (compared to the lowest tertile) was 1.30 (95% CI 1.01, 1.75) per tertile increase of delta biological age. We compared this in the same sample for which our biological aging biomarkers are computed to per tertile change in chronological age 1.90 (95% 1.48, 2.66). BA exceeded CA for prediction of IC (AUC:0.63 vs 0.62; Harrel’s C: 0.68 vs 0.67) (Tables 1-2, Figure 1, Supplementary figures 2-4, Supplementary table 1).

**Table 2.**
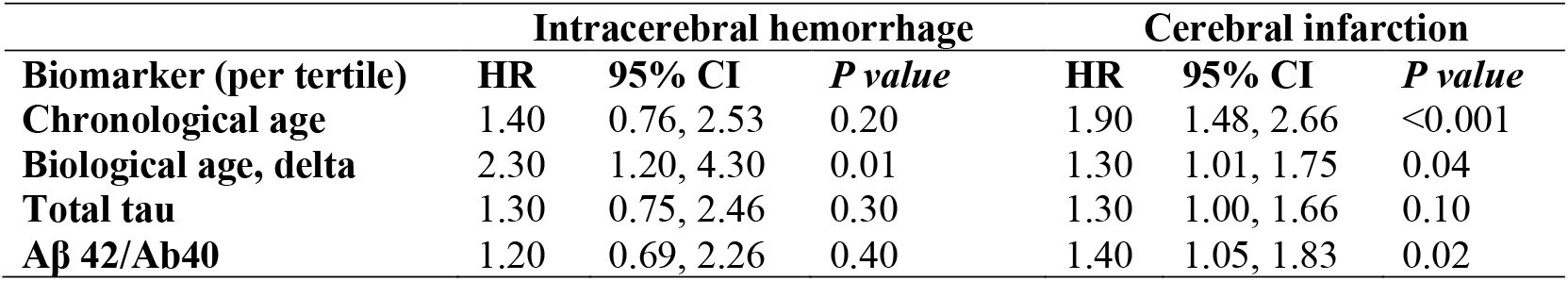
Chronological age, delta biological age, Tau, Aβ 42/Aβ 40 ratio and risk of intracerebral hemorrhage or cerebral infarction.

**Table 3.** Receiver Operating Characteristic (ROC) curve analysis of chronological age and biomarkers as predictors of intracerebral hemorrhage and cerebral infarction.

|  | **Intracerebral hemorrhage** |  | **Cerebral infarction** |  |
| --- | --- | --- | --- | --- |
|  | AUC* | Harrel's C | AUC* | Harrel's C |
| Chronological age | 0.53 (0.41, 0.65) | 0.53 | 0.62 (0.57, 0.67) | 0.66 |
| Biological age | 0.60 (0.49, 0.71) | 0.74 | 0.63 (0.58, 0.67) | 0.67 |
| Delta biological age | 0.68 (0.54, 0.81) | 0.72 | 0.54 (0.46, 0.61) | 0.68 |
| Tau | 0.50 (0.34, 0.66) | 0.56 | 0.55 (0.48, 0.62) | 0.57 |
| Aβ 40 | 0.45 (0.30, 0.60) | 0.52 | 0.55 (0.49, 0.62) | 0.56 |
| Aβ 42 | 0.54 (0.39, 0.69) | 0.58 | 0.59 (0.53, 0.66) | 0.60 |
| Aβ 40/42 ratio | 0.46 (0.32, 0.60) | 0.55 | 0.44 (0.36, 0.50) | 0.58 |
| *Statistical difference between groups: P = 0.18 for intracerebral hemorrhage, P = 0.06 for cerebral infarction |  |  |  |  |

## DISCUSSION

Biological age measured before clinical disease manifestation was able to identify individuals at risk of developing first-ever onset of intracerebral hemorrhage and cerebral infarction with varying magnitude and precision.

The ability of delta biological age, calculated for stroke-free individuals at baseline, to predict risk of intracerebral haemorrhage and cerebral infarction many years before onset could be discussed in several ways. First, the algorithm is based on physiological parameters that reflect the various body systems and the overall underlying individual specific physiological integrity, the various pathophysiological processes and susceptibility to ischemia ^22^. Second, the algorithm was free of brain age biomarkers that are usually significantly higher in later stages along the brain aging continuum and thus offer less opportunities for reversing the progression of cerebrovascular diseases. Furthermore, in our analysis biological aging differences showed varying degrees for risk prediction of cerebral infarction compared to Tau and Beta-amyloid biomarkers^23^. Third, the long follow-up and rigorous clinical examinations allowed precise monitoring of individuals in healthy states prior to clinical manifestation of cerebrovascular disease including strokes. Lastly, the use of the delta biological age measured as the difference between biological age and chronological age reflects within individual variation in aging that cannot be captured by variables measured at time of assessment that mostly reflect cross-sectional data.

Our study has several limitations. First, repeatedly measured biomarker data was not available to calculate that pace of aging that could further confirm the rate of change in biological age. Second, biomarkers of whole blood samples (as HBA1c) were not available; however, the results are comparable as we validated nine versus ten biomarkers in NHANES. Lastly, despite the limited power of the outcome events in the intracerebral hemorrhage group, our sample at baseline was sufficiently powered to detect effect estimates in both groups.

Biological aging is rarely addressed in neurological investigations and its relation to stroke subtypes has not been assessed before. Biological aging is quantified through algorithmic combinations of biological parameters that are sensitive to aging ^17, 24^. Among these approaches, combinations of physiological biomarkers are accessible from blood tests and show minimum biases in relation to method of measurement ^25^. Therefore, the present investigation has important clinical implications. Given the frequency of stroke occurrence and the multi-factorial nature of the disease, development of tools that are accessible and easy to administer in the general practice setting, would accelerate primary prevention efforts of cerebrovascular disorders, through identifying individuals at high risk many years before disease onset. This would offer opportunities for early interventions for those at most risk and potentially reverse risk profiles before clinical manifestation has occurred ^26^. The discovery of minimally invasive and cost-effective biomarkers for cerebrovascular disorders that are extracted from accessible tissue such as serum and sensitive to within individual differences in aging, have the potential to transform clinical research and practice by permitting widespread low-cost screening, risk prediction, and efficient identification of persons with the greatest disease risk for inclusion in clinical trials.

Biological age predicts risk of risk of first-ever ICH and CI in advanced age. The time lag between degradation at the physiological level and onset of cerebrovascular disorders observed in our analysis is an ideal time for intervention in populations at risk. Future studies should confirm our observations and further characterize aging mechanisms that contribute towards brain reserve and resilience among individuals with similar risk profiles.

## Data Availability

Requests to access the dataset from qualified researchers trained in human subject confidentiality protocols may be sent to Department of Epidemiology, Erasmus MC University Medical Center at.

## CONFLICT OF INTEREST

The authors declare that they have no conflicts of interest.

## Figure legends

**Supplementary figure 1.**
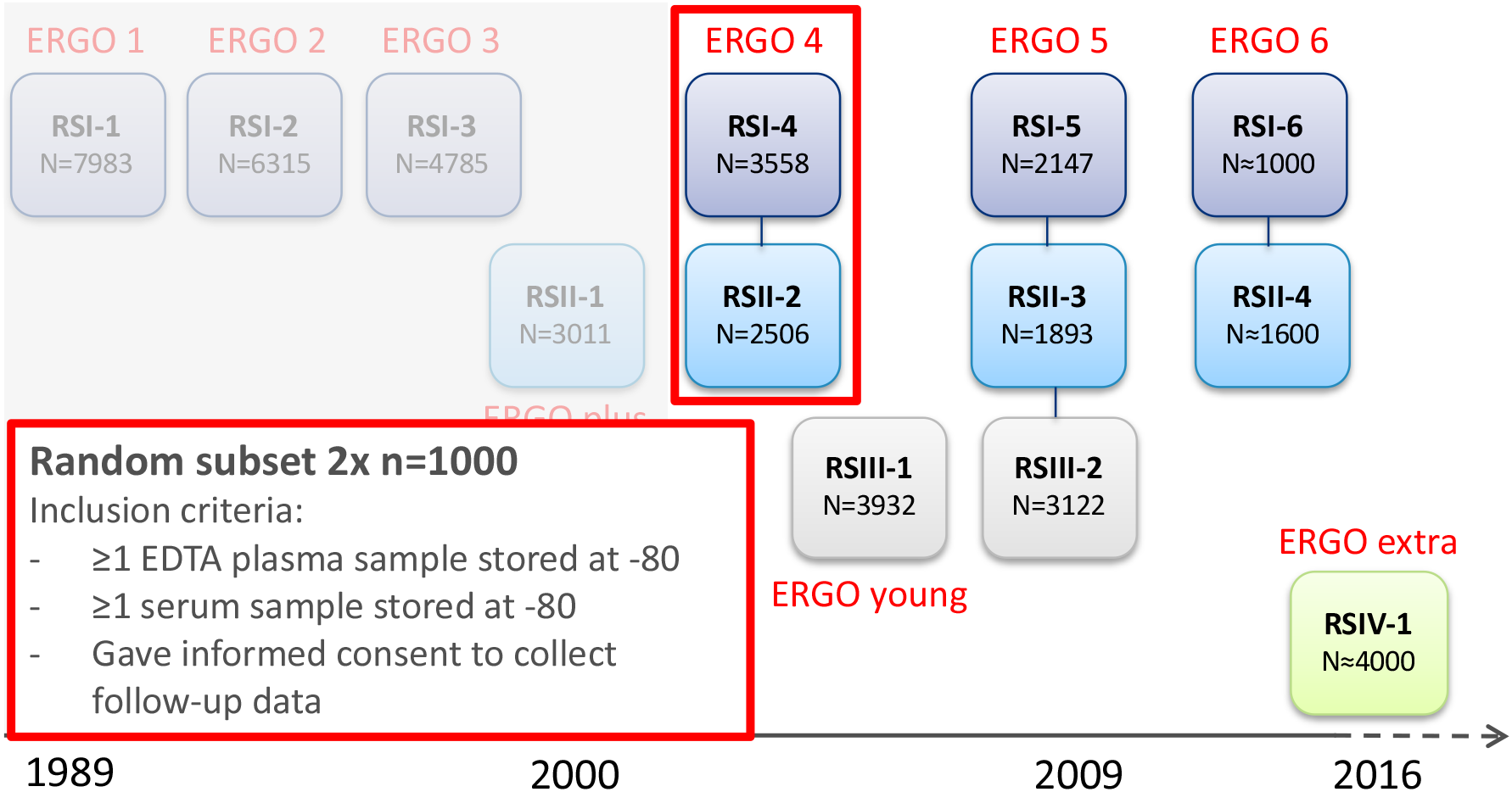
Study design.

**Supplementary Figure 2.**
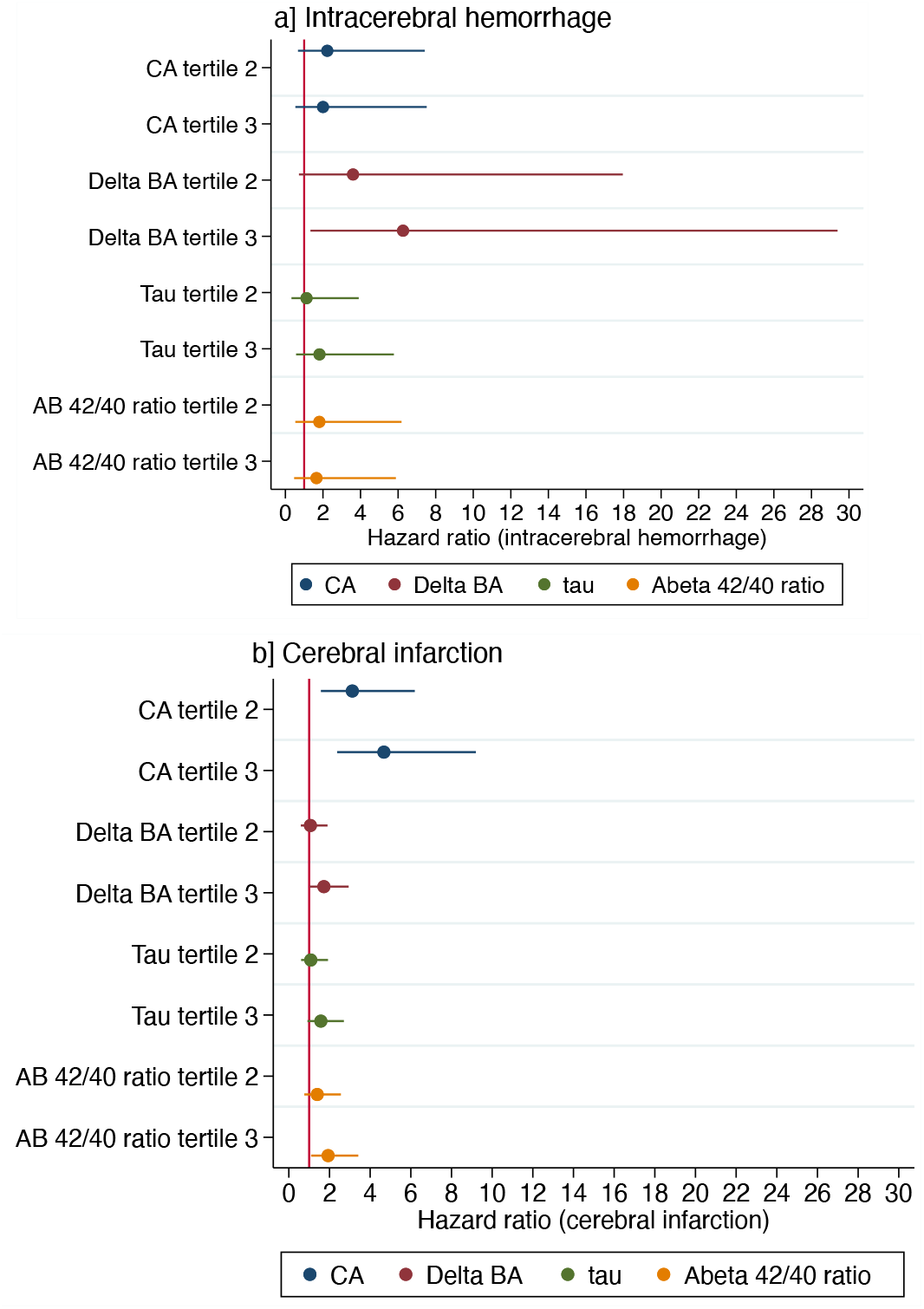
Tertiles of chronological age, delta biological age, Tau, Aβ 42/Aβ 40 ratio as predictors of intracerebral hemorrhage or cerebral infarction.

**Supplementary Figure 3.**
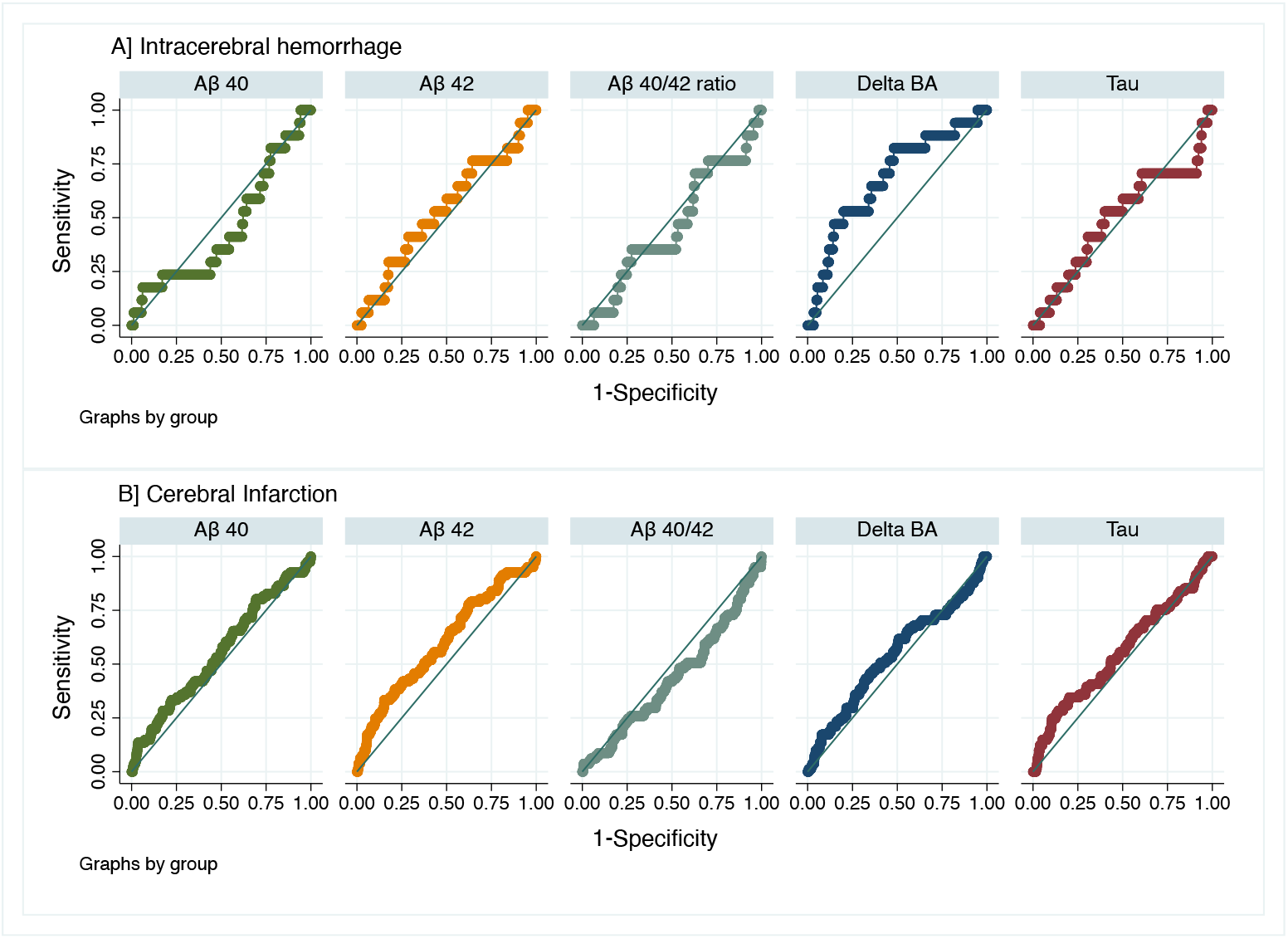
Area under the receiver operating characteristic curve of biomarkers as predictors of a] intracerebral hemorrhage and b] cerebral infarction.

**Supplementary figure 4.**
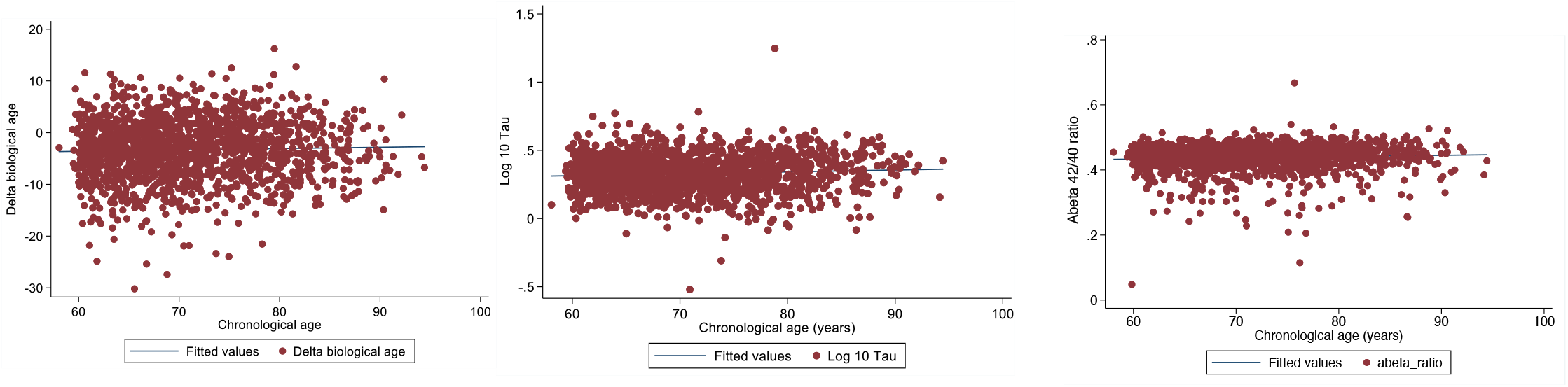
Scatter plots of the correlation between chronological age and biomarkers.

**Supplementary figure 5.**
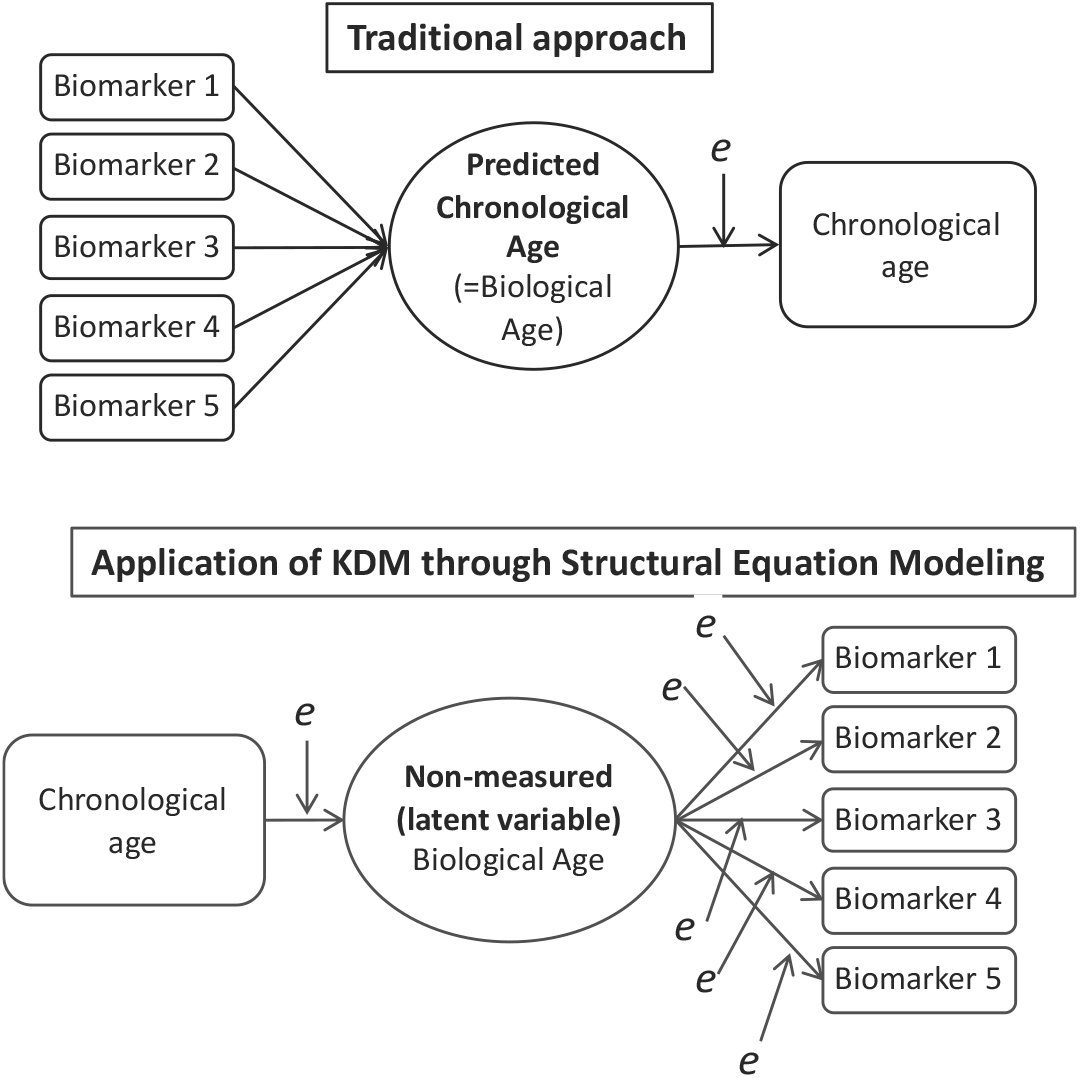
Biological age quantification using structural equation model. Squares: observed | Circles: latent | Arrows: regression coefficients | Arrow points to indicators| E: error Supplementary figure 3. Biological age is a latent intermediate between chronological age and the biomarkers, as opposed to the traditional approach of including chronological age as a dependent variable. Biomarkers are responses to biological age. KDM uses only regression functions of individual biomarkers that can be interpreted as functions of biological age as well as functions of chronological age.

**Supplementary table 1.**
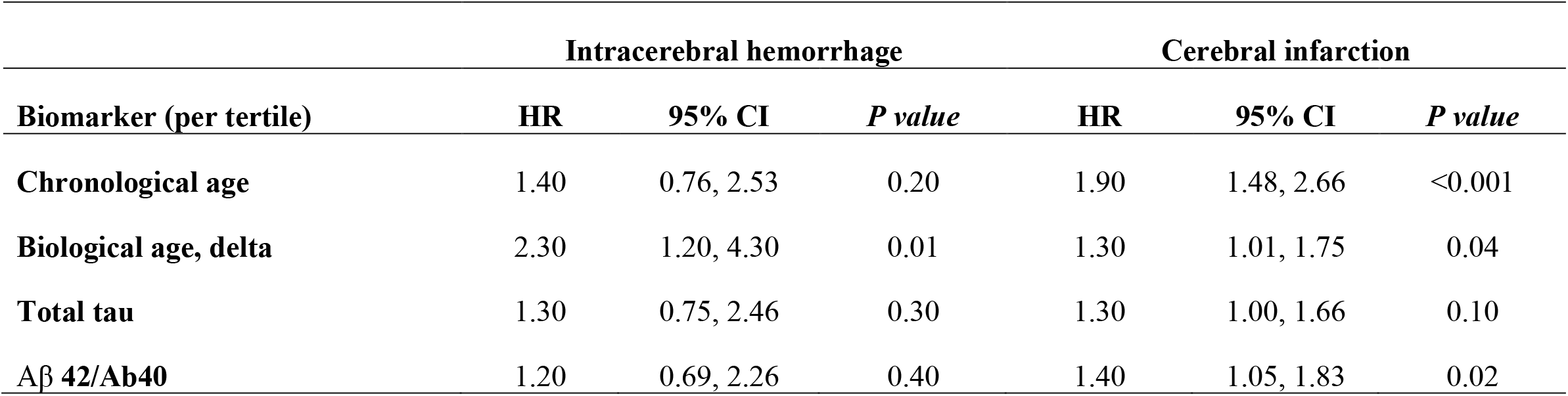
Chronological age, delta biological age, Tau, Aβ 42/Aβ 40 ratio and risk of intracerebral hemorrhage or cerebral infarction.

**Supplementary table 2.** Relative weights of components used in biological age estimation based on NHANES III.

| Parameters | Men |  | Women |  |
| --- | --- | --- | --- | --- |
|  | Original algorithm (10) | Modified algorithm (9) | Original algorithm (10) | Modified algorithm (9) |
| Albumin | 5.8 | 5 | 1.0 | 1 |
| Alkaline phosphatase | 0.8 | 1 | 3.7 | 3 |
| Urea nitrogen | 2.8 | 3 | 5.1 | 5 |
| Creatinine | 2.8 | 3 | 3.1 | 3 |
| CMV | 1.3 | 3 | 0.9 | 2 |
| C-reactive protein | 5.7 | 5 | 2.8 | 3 |
| FEV1 | 19.2 | 15 | 20.6 | 13 |
| HbA1C | 5.2 | na | 5.6 | na |
| Total cholesterol | 4.3 | 4 | 6.0 | 6 |
| Systolic BP | 8.6 | 9 | 12.3 | 13 |
| Age | 43.6 | 51 | 38.9 | 50 |

**Supplementary table 3.** Models fit based on AIC and BIC.

|  | Intracerebral hemorrhage |  | Cerebral infarction |  |
| --- | --- | --- | --- | --- |
|  | AIC | BIC | AIC | BIC |
| CA | 246 | 267 | 1136 | 1158 |
| Delta BA | 242 | 269 | 1135 | 1162 |

**Supplementary table 4.** Tertiles of chronological age, delta biological age, Tau, Aβ 40, Aβ 42 and risk of intracerebral hemorrhage or cerebral infarction with additional APOE4 adjustments.

| **Biomarker (per tertile)** | **Intracerebral hemorrhage** |  |  | **Cerebral infarction** |  |  |
| --- | --- | --- | --- | --- | --- | --- |
|  | **HR<sup>b</sup>** | **95% CI** | ***P value*** | **HR<sup>b</sup>** | **95% CI** | ***P value*** |
| **Chronological age** | 1.40 | 0.76, 2.60 | 0.20 | 1.90 | 1.43, 2.61 | <0.001 |
| **Biological age, delta** | 2.30 | 1.18, 4.55 | 0.01 | 1.30 | 1.00, 1.74 | 0.05 |
| **Tau** | 1.50 | 0.81, 2.80 | 0.10 | 1.30 | 1.02, 1.82 | 0.03 |
| **Aβ 42/Aβ 40** | 1.30 | 0.69, 2.39 | 0.40 | 1.40 | 1.08, 1.95 | 0.01 |

## Notes

### Competing Interest Statement

The authors have declared no competing interest.

### Funding Statement

This study was partially supported by Janssen Prevention Center

## REFERENCES

1. Feigin VL, Norrving B, Mensah GA. Global burden of stroke. Circulation research. 2017;120:439–448

2. Feigin VL, Roth GA, Naghavi M, Parmar P, Krishnamurthi R, Chugh S, et al. Global burden of stroke and risk factors in 188 countries, during 1990–2013: A systematic analysis for the global burden of disease study 2013. The Lancet Neurology. 2016;15:913–924

3. Bennett D, Schneider J, Arvanitakis Z, Kelly J, Aggarwal N, Shah R, et al. Neuropathology of older persons without cognitive impairment from two community-based studies. Neurology. 2006;66:1837–1844

4. Riley KP, Snowdon DA, Markesbery WR. Alzheimer’s neurofibrillary pathology and the spectrum of cognitive function: Findings from the nun study. Annals of neurology. 2002;51:567–577

5. López-Otín C, Blasco MA, Partridge L, Serrano M, Kroemer G. The hallmarks of aging. Cell. 2013;153:1194–1217

6. Hayflick L. The future of ageing. Nature. 2000;408:267–269

7. Sebastiani P, Thyagarajan B, Sun F, Schupf N, Newman AB, Montano M, et al. Biomarker signatures of aging. Aging cell. 2017;16:329–338

8. Chen BH, Marioni RE, Colicino E, Peters MJ, Ward-Caviness CK, Tsai P-C, et al. DNA methylation-based measures of biological age: Meta-analysis predicting time to death. Aging (Albany NY). 2016;8:1844

9. Horvath S, Gurven M, Levine ME, Trumble BC, Kaplan H, Allayee H, et al. An epigenetic clock analysis of race/ethnicity, sex, and coronary heart disease. Genome biology. 2016;17:1–23

10. Levine ME, Lu AT, Bennett DA, Horvath S. Epigenetic age of the pre-frontal cortex is associated with neuritic plaques, amyloid load, and alzheimer’s disease related cognitive functioning. Aging (Albany NY). 2015;7:1198

11. Ikram MA, Brusselle GG, Murad SD, van Duijn CM, Franco OH, Goedegebure A, et al. The rotterdam study: 2018 update on objectives, design and main results. European journal of epidemiology. 2017;32:807–850

12. Ikram MA, Brusselle G, Ghanbari M, Goedegebure A, Ikram MK, Kavousi M, et al. Objectives, design and main findings until 2020 from the rotterdam study. Eur J Epidemiol. 2020;35:483–517

13. Control CfD, Prevention. National center for health statistics (nchs). National health and nutrition examination survey questionnaire (or examination protocol, or laboratory protocol). http://www.cdc.gov/nchs/nhanes.htm. 2006

14. Hatano S. Experience from a multicentre stroke register: A preliminary report. Bull World Health Organ. 1976;54:541–553

15. Wieberdink RG, Ikram MA, Hofman A, Koudstaal PJ, Breteler MM. Trends in stroke incidence rates and stroke risk factors in rotterdam, the netherlands from 1990 to 2008. Eur J Epidemiol. 2012;27:287–295

16. Waziry R, Gras L, Sedaghat S, Tiemeier H, Weverling GJ, Ghanbari M, et al. Quantification of biological age as a determinant of age-related diseases in the rotterdam study: A structural equation modeling approach. European journal of epidemiology. 2019;34:793–799

17. Levine ME. Modeling the rate of senescence: Can estimated biological age predict mortality more accurately than chronological age? Journals of Gerontology Series A: Biomedical Sciences and Medical Sciences. 2013;68:667–674

18. Belsky DW, Caspi A, Houts R, Cohen HJ, Corcoran DL, Danese A, et al. Quantification of biological aging in young adults. Proceedings of the National Academy of Sciences. 2015;112:E4104–E4110

19. Holtzman S, Vezzu S. Confirmatory factor analysis and structural equation modeling of noncognitive assessments using proc calis. NorthEast SAS Users Group (NESUG), 2011 proceedings. 2011:11–14

20. Jöreskog KG. A general method for analysis of covariance structures. Biometrika. 1970;57:239–251

21. Waziry R, Heshmatollah A, Bos D, Chibnik LB, Ikram MA, Hofman A, et al. Time trends in survival following first hemorrhagic or ischemic stroke between 1991 and 2015 in the rotterdam study. Stroke. 2020;51:824–829

22. Jylhävä J, Pedersen NL, Hägg S. Biological age predictors. EBioMedicine. 2017;21:29–36

23. Vemuri P, Lesnick TG, Knopman DS, Przybelski SA, Reid RI, Mielke MM, et al. Amyloid, vascular, and resilience pathways associated with cognitive aging. Annals of neurology. 2019;86:866–877

24. Liu Z, Kuo P-L, Horvath S, Crimmins E, Ferrucci L, Levine M. A new aging measure captures morbidity and mortality risk across diverse subpopulations from nhanes iv: A cohort study. PLoS medicine. 2018;15:e1002718

25. Levine ME. Response to dr. Mitnitski’s and dr. Rockwood’s letter to the editor: Biological age revisited. Journals of Gerontology Series A: Biomedical Sciences and Medical Sciences. 2014;69:297–298

26. Balami JS, Buchan AM. Complications of intracerebral haemorrhage. The Lancet Neurology. 2012;11:101–118

27. Heshmatollah A, Fani L, Koudstaal PJ, Ghanbari M, Ikram MA, Ikram MK. Plasma amyloid beta, total-tau and neurofilament light chain levels and the risk of stroke: A prospective population-based study. Submitted April 2021.

